# Costs and cost-effectiveness of an infection prevention bundle to reduce neonatal sepsis and mortality in Zambia: The Sepsis Prevention in Neonates in Zambia (SPINZ) trial

**DOI:** 10.64898/2026.02.04.26345617

**Authors:** Lora L Sabin, Rebecca L West, Susan E. Coffin, Sylvia Machona, Carter Cowden, Lawrence Mwananyanda, Chileshe Lukwesa-Musyani, John Tembo, Matthew Bates, Davidson H. Hamer

## Abstract

The Sepsis Prevention in Neonates in Zambia (SPINZ) trial was a prospective observational cohort study conducted in the neonatal intensive care unit of the University Teaching Hospital in Lusaka, Zambia. Introduction of an infection prevention and control (IPC) bundle reduced hospital-associated mortality, total mortality, suspected sepsis, and confirmed bloodstream infections. This companion analysis was undertaken to analyze intervention costs and cost-effectiveness in this low-resource setting. We conducted a retrospective cost analysis, using SPINZ study-related records, and expressed costs in real 2016 US dollars. We also estimated intervention cost-effectiveness using both outcomes from SPINZ (avoided deaths, confirmed infections, and suspected episodes of infection) and estimated disability-adjusted life years (DALYs) averted by the intervention. To provide data for policymakers, a future cost projection was undertaken to estimate costs of the program implemented nationally over a 10-year period in real 2025 US dollars. A total of 2,035 neonates were enrolled from September 2015 to March 2017. Total costs during implementation (introduction of the IPC bundle) (April-May 2016) and the subsequent intervention period were $17,641 and $5,265, respectively, of which most expenses were incurred during the preparation period due to travel and training. During the intervention period, the program’s running cost was approximately $478 per month. The estimated cost per death, confirmed infection, and suspected episode averted was $208, $204, and $32, respectively; the estimated cost per DALY averted was $9.5. The future model was estimated to cost an average of $107,561 annually to implement nationally. The analysis indicated that the IPC bundle to prevent sepsis-related neonatal mortality was highly cost-effective. Cost reductions from task-shifting, reduced preparation (start-up) costs, and longer intervention periods would further decrease cost per death averted. IPC bundle implementation can thus be recommended for resource-constrained settings where sepsis and other nosocomial infections are associated with high neonatal mortality.

## Introduction

Nearly half of all under-5 deaths occur in the 1st 28 days of life [1, 2]. Thus, preventing deaths in this vulnerable age group would contribute to a significant reduction in under-five mortality rates globally. Neonatal sepsis, a leading cause of morbidity and mortality among newborn infants worldwide, affects approximately three million newborn infants and results in 11-19% mortality [3]. The World Health Organization (WHO) has declared sepsis a global priority due to this impact [4, 5]. The burden of neonatal sepsis is particularly high in low- and middle-income countries (LMICs) [6], where incidence is as high as 49 to 170 cases per 1,000 live births, with a case fatality rate of up to 24% [3,4].

In 2022, the WHO reported the highest neonatal mortality rate in sub-Saharan Africa, where there were 27 deaths per 1,000 live births across the region [7]. This may be partly an unintended consequence of efforts to increase the number of facility-based deliveries in sub-Saharan countries since 2010; one the most common adverse events arising from a hospital-based delivery is a higher risk of nosocomial (hospital-acquired) infections [8, 9]. This risk is particularly important given evidence that nearly 40% of newborn deaths in LMICs are linked to nosocomial infections [7]. The most common nosocomial cause of neonatal deaths is bloodstream infection (BSI), with gram-negative bacteria being the most common causative organism of neonatal sepsis [10, 11].

Despite making significant improvements to improve infant health outcomes in Zambia in recent decades, Zambia still performs poorly relative to other countries due to its neonatal mortality rate, ranking 162 out of 195 countries [12]. An estimated 1 in 37 Zambian infants die in their first month of life [13]. To address this situation, a prospective observational cohort study, the Sepsis Prevention in Neonates in Zambia (SPINZ) study, was conducted between 2015 and 2017 to test the impact of an infection prevention and control (IPC) bundle on BSI and sepsis-induced neonatal mortality [14]. The IPC bundle was associated with significantly reduced BSI and mortality, providing important evidence on an approach to reduce neonatal mortality in resource-constrained regions (see study findings reported previously) [14]. Given these promising results, we conducted a supplemental cost analysis. The analysis included a retrospective cost and cost-effectiveness analysis of the SPINZ study, as well as a forward-looking estimate of financial costs for broader IPC bundle implementation over a 10-year timeframe. The goal of this analysis is to provide policy-relevant information for potential scale-up of the intervention package in Zambia or other resource-limited countries for prevention of neonatal sepsis and reduction of sepsis-associated neonatal mortality.

## Materials and Methods

### SPINZ study summary

SPINZ was a prospective observational cohort study conducted in the neonatal intensive care unit (NICU) of the University Teaching Hospital (UTH) in Lusaka, Zambia between September 2015 and March 2017 [14]. A total of 2,035 neonates admitted to the NICU on weekdays between 8 AM and 5 PM were enrolled. A multifaceted IPC bundle was implemented that consisted of 1) training of all NICU staff (physicians, nurses, pharmacists, and environmental health technologists) on the theory and practice of measures to prevent nosocomial infections with an emphasis on proper hand hygiene; 2) daily text message reminders on IPC; 3) alcohol hand rub; 4) enhanced environmental cleaning; and 5) weekly bathing of babies with 2% chlorhexidine gluconate. The primary study outcome was hospital-associated mortality among enrolled neonates in the UTH NICU. Secondary outcomes were incidence of hospital-associated suspected sepsis and BSI with pathogens. The study timeline included an initial six-month baseline period from September 2015 to March 2016; six weeks to implement the IPC bundle intervention, including training from April to May 2016; and 11 months of intervention with assessment from June 2016 to March 2017. A time series analysis of the SPINZ study interventions revealed reductions in hospital-associated mortality, total mortality, suspected sepsis, and confirmed BSI with pathogen [14].

The study was approved by the ethical committees at Boston University Medical Campus, Excellence in Research Ethics and Science Converge in Zambia, and the Children’s Hospital of Philadelphia, as well as the Zambian Ministry of Health Research Secretariat and the UTH Department of Pediatrics. Written informed consent was provided by the participating neonate’s mother or legal guardian either in English or the local languages (Nyanja and Bemba).

### Cost analyses

Two financial analyses were conducted. The first focused on the financial costs incurred during the SPINZ trial itself, encompassing the approximately 13-month implementation and intervention/assessment periods (excluding the baseline period). To provide information for policymakers on the cost of major intervention scale-up [15], a second forecasted analysis modeled the expected cost to deliver the intervention nationwide over a future 10-year timeframe from the provider’s perspective.

#### Financial cost of the intervention

The financial analysis of the trial was based on actual expenditures related to the SPINZ implementation and intervention/assessment periods. This included expenditures associated with both the implementation phase from April to May 2016, and 11 months of intervention and assessment from June 2016 through April 2017. Cost data were collected retrospectively from program documents and invoices of payments made over the study period. Estimates of costs were made for printing training materials and posters, promotional lapel pins during training, volume measuring containers for alcohol-based hand rub production, chlorhexidine wipes, chlorhexidine shipping, plastic baby dolls for training, and bleach for sink cleaning, because precise documentation was not available.

Costs were organized by local (Zambia) and US-based expenses over the entire 13-month period and then categorized (Table 1). Costs were also organized by project period (implementation, e.g. preparation) and intervention/assessment). We also categorized fixed costs unrelated to the number of participating nurses (e.g., travel, training, and some supplies) versus variable costs (SMS and other supplies). During the intervention/assessment period, clinician salaries were not covered by study funds. In contrast, a portion of the NICU pharmacist’s salary was included in personnel costs during both the implementation and intervention/assessment periods and an incentive for the nursing staff was included in personnel costs but only during the intervention/assessment periods. All research costs were excluded. Total nominal Zambia-based costs were converted into US dollar equivalents using a mean exchange rate of ZMK 10.31 for 2016 and ZMK 9.52 for 2017 and added to total US dollar costs. We then adjusted total annual US dollar costs by US inflation rates (Consumer Price Index) to calculate costs in real 2016 US dollars. Finally, we applied a discount rate of 3% to 2017 costs to estimate the total 2016 present value of intervention delivery.

**Table 1.**
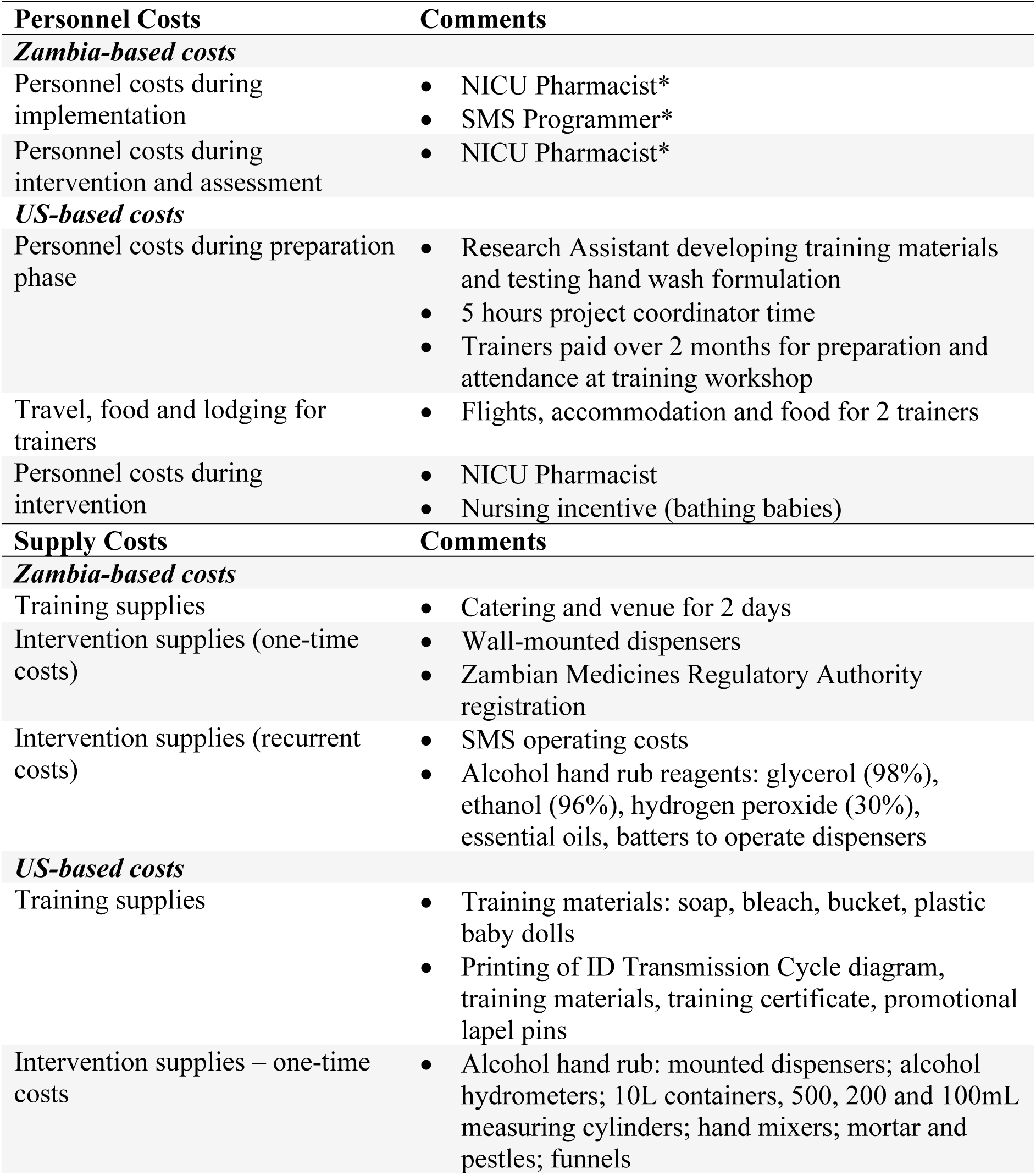
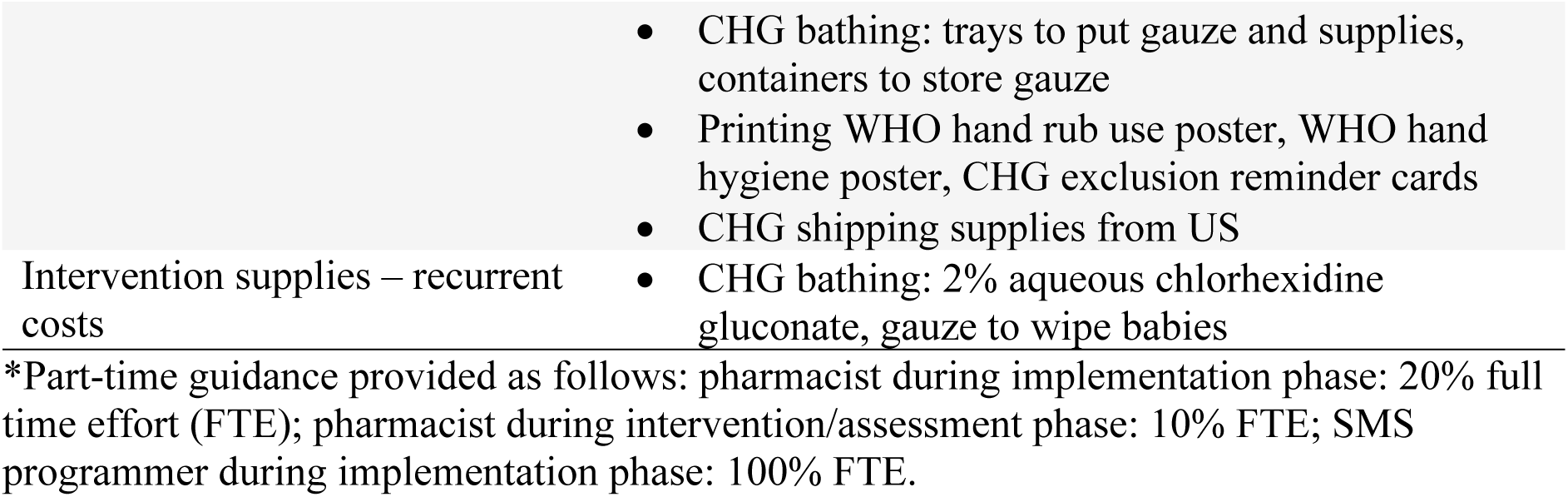
Cost categories and details: SPINZ implementation and intervention/assessment.

#### Forecasted 10-year cost analysis

We utilized a financial perspective to estimate program delivery over a future 10-year lifespan from 2025-2035 to be scaled up to all tertiary and district hospitals in Zambia. We assumed a similar model as that of the trial but added a month to the preparation period to be conservative. Thus, the timeline encompassed 3 months for preparation followed by 9 years and 9 months of intervention implementation.

In the forecasted model, 1 cost unit was applied for tertiary hospitals and 0.5 cost units were applied for district hospitals, using an estimate of 6 tertiary hospitals and 29 district (second-level) hospitals nationally [16]. Where possible, costs for salaries and supplies were provided by the Zambian team; otherwise, estimates collected for the original analysis were used and adjusted for inflation at a rate of 8.02%. Adjustments were made to the model to increase local sustainability: all staff were based in Zambia (e.g. no U.S.-based personnel were included), supplies were all purchased in Zambia, and non-essential supplies (such as essential oils for hand rub solution) or one-off costs for the SPINZ study that did not apply in this scenario (e.g. Zambia Medicines Regulatory Authority [ZAMRA] registration) were excluded. Nurse incentives for bathing babies were removed as the program was assumed to be integrated into standard care.

The Zambian-based NICU consultant was excluded as this role would not be relevant in a scale-up scenario. Annual refresher training costs were included to account for staff attrition and need for ongoing medical education. Annual Zambia-based costs in local currency were converted to nominal US dollars using an expected constant real exchange rate equal to the average 2025 rate of 28.1 ZMK = 1 USD. Total annual costs were adjusted for expected inflation [17] and discounted at a rate of 3% annually per economic evaluation best practice [18].

Given the uncertainty in estimating costs so broadly and over a 10 year life span, sensitivity analyses were also conducted utilizing two scenarios which adjusted costs by 25% upward and downward, respectively: 1) a maximum cost scenario where a tertiary hospital = 1.25 cost units and a second-level hospital = 0.75 cost units and 2) a minimum cost scenario where a tertiary hospital = 0.75 cost units and a second-level hospital = 0.25 cost units.

### Cost-effectiveness analyses

The cost-effectiveness of the SPINZ intervention utilized an adapted version of the standard formula: *ICEA = (C_I_ - C_C_)/(O_I_ -O_C_)* where ICEA is the incremental cost-effectiveness ratio. Typically, C_I_ and C_C_ are the total (discounted) costs associated with intervention and control or comparison groups, respectively. In this analysis, all costs were incremental and there was no control group, so C_C_ = 0. For C_I_, we utilized the result of the financial cost analysis described above. Typically, O_I_, O_C_ are the (discounted) morbidity or mortality outcomes for intervention and control groups, respectively. In this analysis, intervention outcomes were taken directly from the trial’s estimated effects, based on pre-post analyses, including avoided deaths, suspected episodes of sepsis averted, and laboratory-confirmed infections (BSI with pathogen) averted. We also estimated the number of averted disability-adjusted life years (DALYs) using an approach that incorporates expected years of life lost (not country-specific) with time lived at different ages using an exponential function and discounted at 3% [19].

For the forecasted CEA, we used the estimated base case economic costs for the 10-year program and utilized the trial’s 17% effect on reduced mortality for both expected mortality and DALYs averted. The number of annual admissions into NICUs was estimated using the mean number of study enrollments during the SPINZ intervention (325 per month * 12 months); units of 1.0 and 0.5 were applied to represent admissions annually to tertiary and secondary hospitals, respectively. A rate of 0.02% was applied to expected increase of admissions annually. In addition, we examined the cost-effectiveness of two alternative ‘optimistic’ and ‘conservative’ scenarios where reduced mortality rates were 25% and 10%, respectively, in the two scenarios.

Interpretations regarding cost-effectiveness were based on the 2001 recommendation of the Commission on Macroeconomics and Health and adopted by the World Health Organization (WHO) whereby an intervention is ‘highly cost-effective’ if it averts a DALY for less than per capita GDP (Gross Domestic Product) and ‘cost-effective’ if it averts a DALY for less than 3 times per capita GDP [20].

## Results

### SPINZ Program costs

In the financial analysis, the total program cost in real discounted 2016 US$ was $22,419, of which $17,641 was incurred in the preparation period (April-May 2016) and $4,778 in the implementation period (Table 2). During the former, training activities accounted for the majority of costs (74%), including venue and food in Zambia, US-based trainer salaries, and trainer flights and accommodation. Supplies comprised approximately 10% of costs, while SMS set-up costs accounted for another 9%. The remaining costs were other direct costs, including training materials, printing, and batteries. During the implementation period, the greatest cost was compensation to the NICU Pharmacist Consultant, which accounted for 42% of costs. The next largest expense was alcohol hand rub, which accounted for just under 25% of costs.

**Table 2.** SPINZ Total Costs by Study Period.

| **Program Period** | **Cost** |
| --- | --- |
| **Implementation (Apr-May 2016)** |  |
| ***Personnel*** |  |
| NICU pharmacist | \$ 388 |
| US-based trainers | \$ 3,616 |
| Research assistant | \$ 600 |
| Project coordinator | \$ 108 |
| ***Fixed Costs*** |  |
| *1. Travel* |  |
| Trainer flights | \$ 4,851 |
| Accommodation and food (trainers) | \$ 2,112 |
| *2. Training* |  |
| Venue and food | \$ 2,328 |
| Training materials and printing promotional buttons | \$ 162 |
| *3. Supplies* |  |
| Wall-mounted dispensers | \$ 436 |
| Soap, water, hand rub solution | \$ 132 |
| Supplies for chlorhexidine bathing | \$ 30 |
| *4. Other direct costs* |  |
| SMS Set-Up | \$ 1,600 |
| Zambia Medicines Regulatory Authority registration for chlorhexidine | \$ 6 |
| Hand rub use and hygiene printing | \$ 2 |
| Batteries for dispensers | \$ 15 |
| Chlorhexidine shipping (supplies) | \$ 100 |
| ***Variable costs*** |  |
| *1. Supplies* |  |
| CHG Bathing | \$ 953 |
| *Sub-Total Preparation, Current US\$* | *\$ 17,641* |
| *Sub-Total Preparation, Real 2016 US\$* | *\$ 17,641* |
| *Sub-Total Preparation, Discounted Real 2016 US\$* | *\$ 17,641* |
| **Intervention/assessment (June 2016 – March 2017)** |  |
| ***Personnel*** |  |
| Nurses (incentives) | \$ 895 |
| NICU Pharmacist | \$ 1,988 |
| ***Fixed Costs*** |  |
| *1. Supplies* |  |
| Alcohol hand rub | \$ 1,152 |
| *2. Other Direct Costs* |  |
| SMS Operating Costs | \$ 819 |
| *Sub-Total Intervention Implementation, Current US\$* | *\$ 4,853* |
| *Sub-Total Intervention Implementation, Real 2016 US\$* | *\$ 4,822* |
| *Sub-Total Intervention Implementation, Discounted Real 2016 US\$* | *\$ 4,778* |
| **Total Costs, Current US\$** | **\$ 22,494** |
| **Total Costs, Real 2016 US\$** | **\$ 22,463** |
| **Total Discounted Costs, Real 2016 US\$** | **\$ 22,419** |

### Forecasted analysis

Total forecasted costs for a 10-year national program in 2025 US$ (real, discounted) were $1,075,613 (Table 3). This equates to $107,561 annually, or $5,246 per year per tertiary hospital (assuming 1.0 cost units per hospital) and $2,623 per second-level hospital (assuming 0.5 cost units per hospital). Intervention costs accounted for 96% of costs overall; 29% were fixed costs (primarily SMS operation during intervention implementation, and annual materials required for dispensing alcohol hand rub). The most significant cost drivers in the forecasted model were variable supply costs for intervention, including alcohol hand rub, CHG bathing, and cleaning (altogether, accounting for 63% of costs).

**Table 3.**
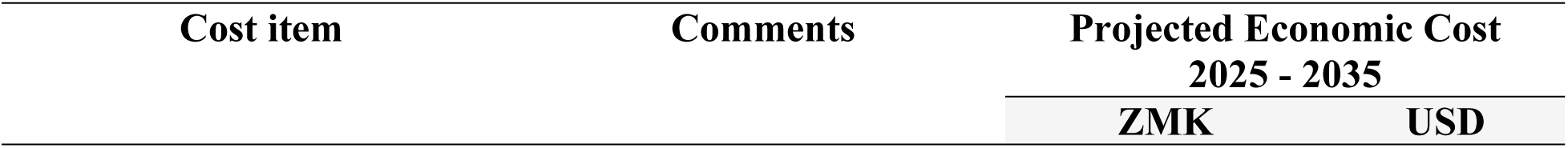
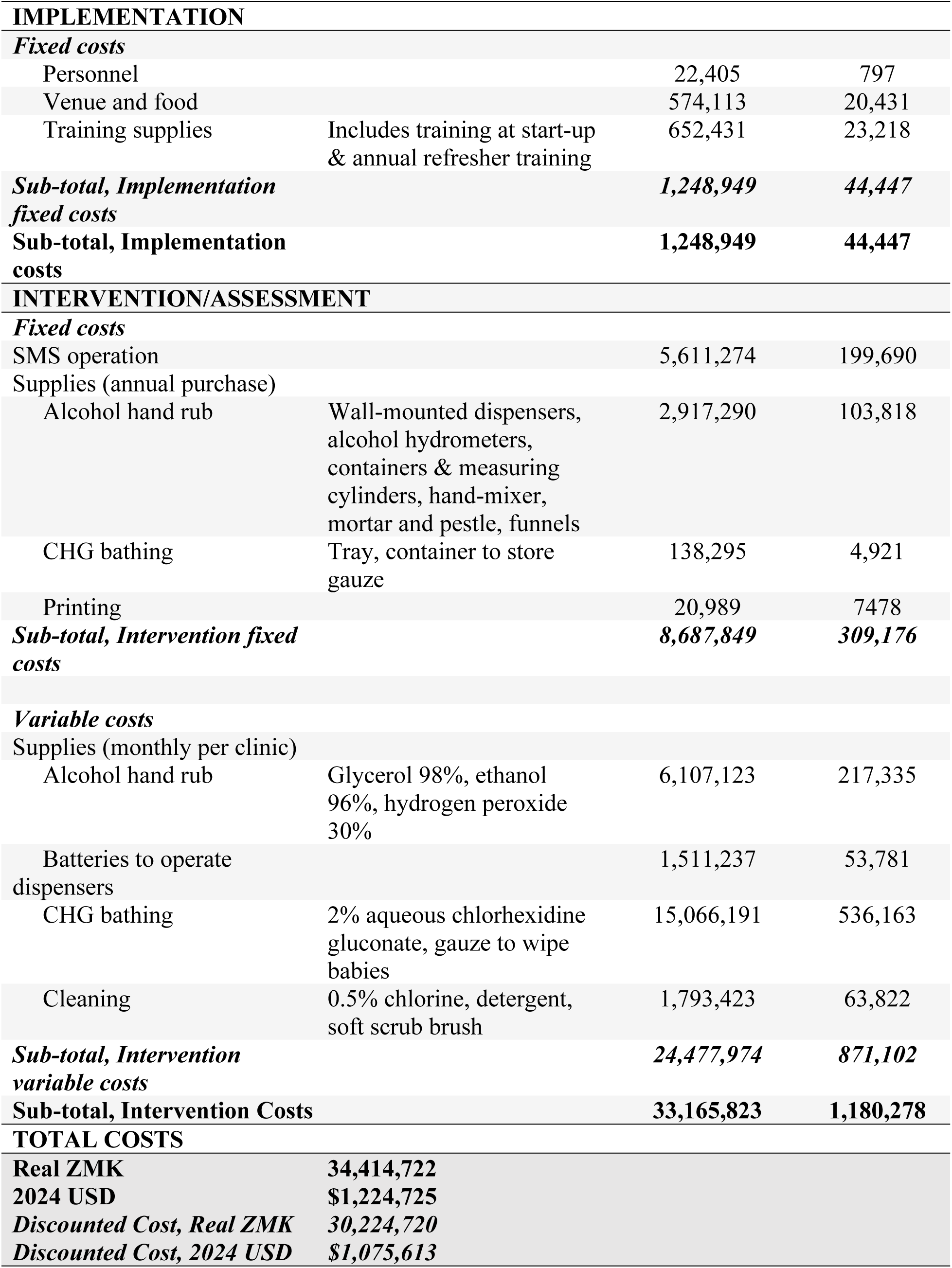
Forecasted analysis, 10-year national program.

Sensitivity analysis of the 10-year scaled-up program found that costs could range from $153,472 per year (in the maximum cost scenario) to $61,650 per year (in the minimum cost scenario (Table 4). In the maximum cost scenario, annual program cost would be $6,559 and $3,935 for a tertiary and second-level hospital, respectively. Alternatively, the minimum cost scenario would be $3,935 and $1,311, respectively, for the two hospital levels.

**Table 4.**
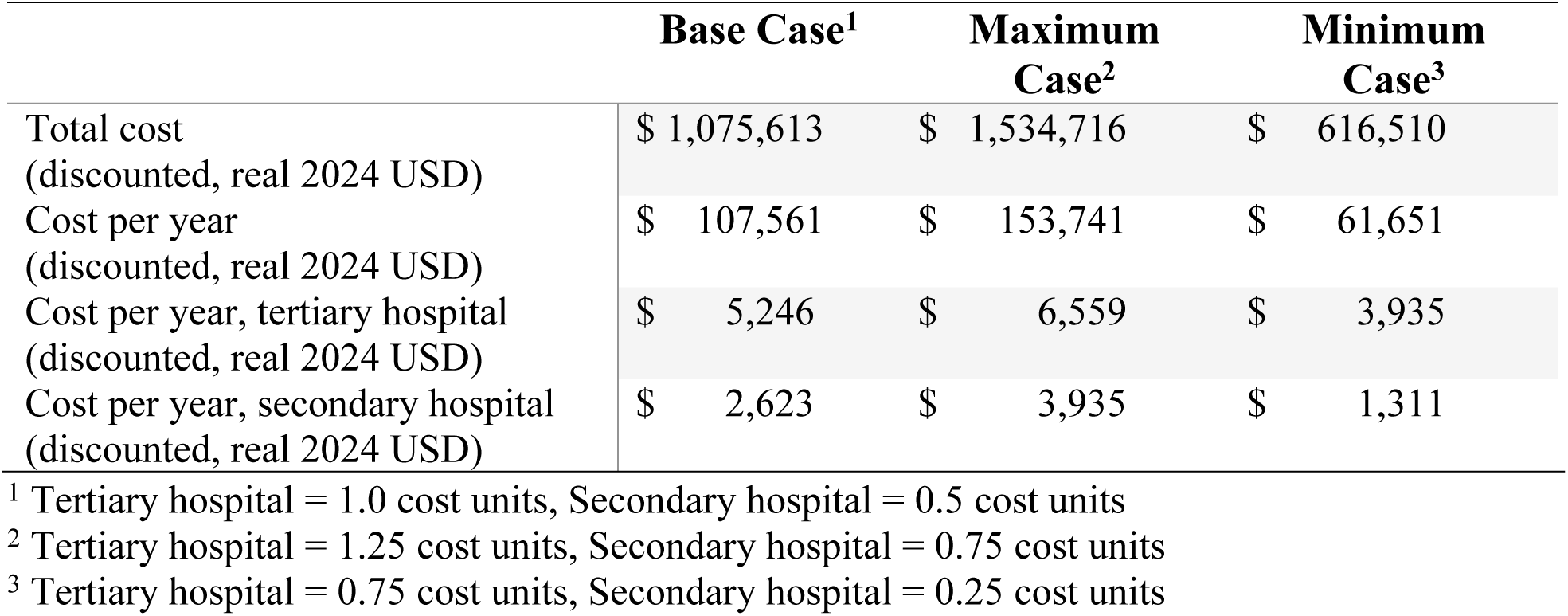
Forecasted program, sensitivity analysis.

### Cost-effectiveness analysis

As shown in Table 5, the SPINZ bundle of IPC interventions resulted in avoidance of 108 neonatal deaths, at a cost of $207.6 per death avoided; 704 suspected episodes of sepsis were averted, at a cost of $31.8 per episode averted. A total of 110 confirmed infections of sepsis were averted, at a cost of $204.2 per infection averted. Additionally, an estimated 21.9 DALYs were averted per neonatal death avoided (total of 2,365 DALYs) at a cost of $9.5 per DALY averted.

**Table 5.**
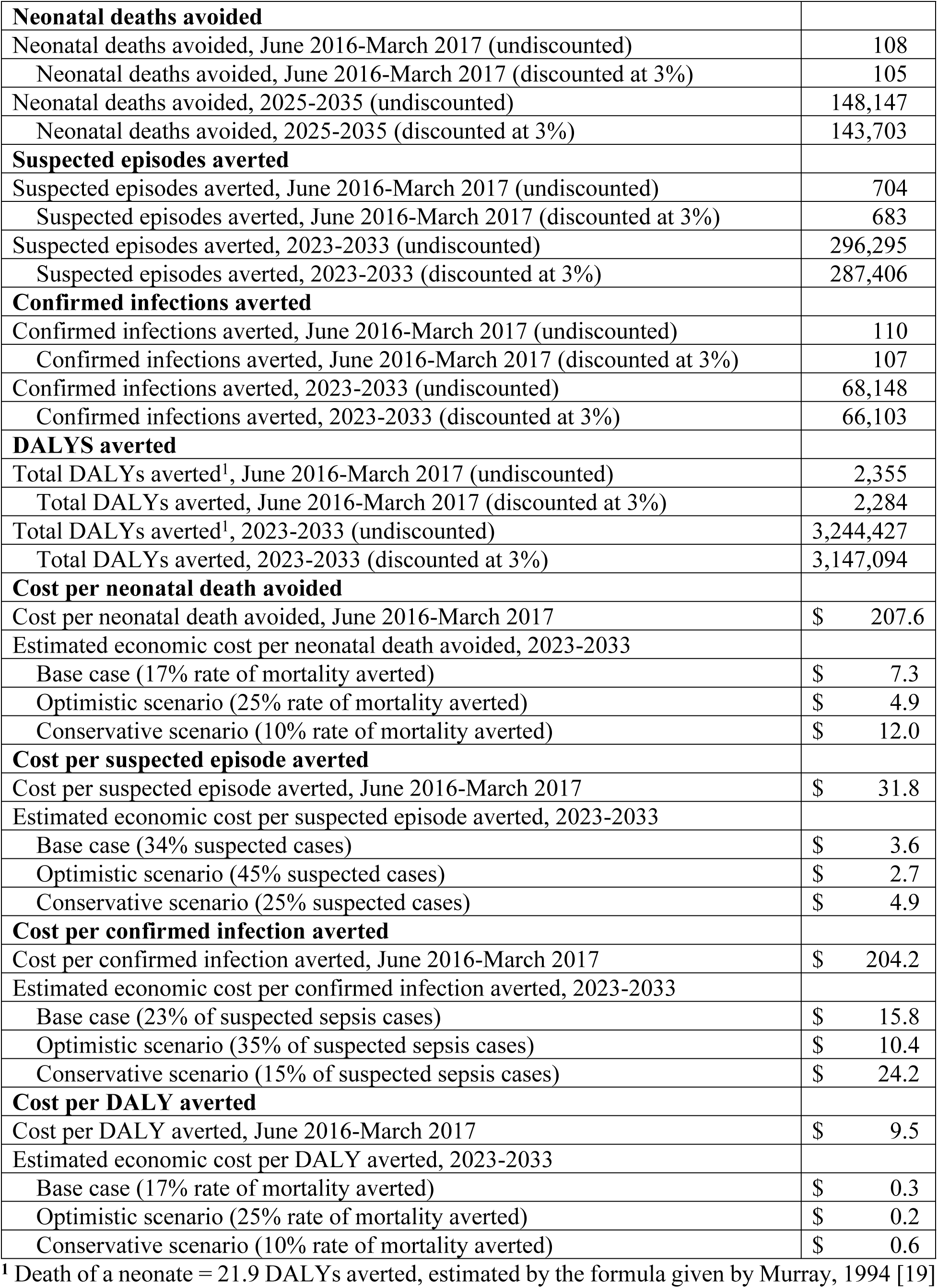
Program cost-effectiveness.

|  |  |
| --- | --- |
| **Neonatal deaths avoided** |  |
| Neonatal deaths avoided, June 2016-March 2017 (undiscounted) | 108 |
| Neonatal deaths avoided, June 2016-March 2017 (discounted at 3%) | 105 |
| Neonatal deaths avoided, 2025-2035 (undiscounted) | 148,147 |
| Neonatal deaths avoided, 2025-2035 (discounted at 3%) | 143,703 |
| **Suspected episodes averted** |  |
| Suspected episodes averted, June 2016-March 2017 (undiscounted) | 704 |
| Suspected episodes averted, June 2016-March 2017 (discounted at 3%) | 683 |
| Suspected episodes averted, 2023-2033 (undiscounted) | 296,295 |
| Suspected episodes averted, 2023-2033 (discounted at 3%) | 287,406 |
| **Confirmed infections averted** |  |
| Confirmed infections averted, June 2016-March 2017 (undiscounted) | 110 |
| Confirmed infections averted, June 2016-March 2017 (discounted at 3%) | 107 |
| Confirmed infections averted, 2023-2033 (undiscounted) | 68,148 |
| Confirmed infections averted, 2023-2033 (discounted at 3%) | 66,103 |
| **DALYS averted** |  |
| Total DALYs averted<sup>1</sup>, June 2016-March 2017 (undiscounted) | 2,355 |
| Total DALYs averted, June 2016-March 2017 (discounted at 3%) | 2,284 |
| Total DALYs averted<sup>1</sup>, 2023-2033 (undiscounted) | 3,244,427 |
| Total DALYs averted, 2023-2033 (discounted at 3%) | 3,147,094 |
| **Cost per neonatal death avoided** |  |
| Cost per neonatal death avoided, June 2016-March 2017 | \$ 207.6 |
| Estimated economic cost per neonatal death avoided, 2023-2033 |  |
| Base case (17% rate of mortality averted) | \$ 7.3 |
| Optimistic scenario (25% rate of mortality averted) | \$ 4.9 |
| Conservative scenario (10% rate of mortality averted) | \$ 12.0 |
| **Cost per suspected episode averted** |  |
| Cost per suspected episode averted, June 2016-March 2017 | \$ 31.8 |
| Estimated economic cost per suspected episode averted, 2023-2033 |  |
| Base case (34% suspected cases) | \$ 3.6 |
| Optimistic scenario (45% suspected cases) | \$ 2.7 |
| Conservative scenario (25% suspected cases) | \$ 4.9 |
| **Cost per confirmed infection averted** |  |
| Cost per confirmed infection averted, June 2016-March 2017 | \$ 204.2 |
| Estimated economic cost per confirmed infection averted, 2023-2033 |  |
| Base case (23% of suspected sepsis cases) | \$ 15.8 |
| Optimistic scenario (35% of suspected sepsis cases) | \$ 10.4 |
| Conservative scenario (15% of suspected sepsis cases) | \$ 24.2 |
| **Cost per DALY averted** |  |
| Cost per DALY averted, June 2016-March 2017 | \$ 9.5 |
| Estimated economic cost per DALY averted, 2023-2033 |  |
| Base case (17% rate of mortality averted) | \$ 0.3 |
| Optimistic scenario (25% rate of mortality averted) | \$ 0.2 |

|  |  |  |
| --- | --- | --- |
| Conservative scenario (10% rate of mortality averted) | \$ | 0.6 |

<sup>1</sup> Death of a neonate = 21.9 DALYs averted, estimated by the formula given by Murray, 1994 [19]

For the ten-year intervention program, an estimated 148,147 neonatal deaths would be avoided at a cost of $7.3 per death avoided. A total of 296,295 suspected episodes of sepsis would be averted at a cost of $3.6 per episode; 68,148 laboratory-confirmed infections of sepsis would be avoided at a cost of $15.8 per infection. Additionally, an estimated 3,244,427 DALYs would be averted at a cost of $0.3 per DALY averted.

## Discussion

The IPC bundle in a NICU in Zambia was effective in preventing suspected sepsis episodes, confirmed BSI, and sepsis-related neonatal mortality, and was a highly cost-effective intervention. Based on our projected analysis, cost reductions from task shifting and longer intervention periods would likely further reduce the cost per death averted. This IPC bundle intervention could thus be recommended in similar low-resource settings where sepsis and nosocomial infections are associated with high neonatal mortality given its cost effectiveness.

A recent comprehensive review highlighted the importance of multimodal interventions for prevention of serious neonatal infections in health facilities [21]. Multi-component interventions involving regulation and restriction reduced the risk of neonatal mortality secondary to BSI by 32% while approaches that included regulation, restriction, and education were associated with reductions in all-cause neonatal mortality by 27%. These interventions were also associated with reductions in antimicrobial resistance. The SPINZ study used a similar multi-component approach with education and epidermal barrier interventions [14]. The SPINZ intervention was designed to be low cost and scalable. The current analysis confirms that this bundle strategy was highly cost effective.

There have been relatively few cost effectiveness analyses of multicomponent facility-level interventions designed to reduce neonatal sepsis and improve newborn survival. One important component of SPINZ was developing local capacity to create alcohol-based hand rub, making the alcohol-based hand rub readily available throughout the NICU, and educating NICU staff about the importance of handwashing. Notably, a NICU-based study in Ghana using a similar pre-post study design showed that the use of alcohol-based hand rub reduced neonatal BSI, BSI-related mortality, BSI-attributable hospital costs, and the patient cost of neonatal BSI [22]. The ideal combination of interventions for reducing nosocomial infections and improving newborn health outcomes may depend on the local context, but better hand hygiene is an important component of multi-component intervention strategies.

While the SPINZ intervention itself was cost-effective, the future forecasted analysis of a nationally scaled up program proved to be even more so due to a number of factors. First, spreading fixed costs across the ten-year period makes costs more manageable for the health system to absorb as well as making the intervention sustainable over the long term. Localizing the program to be run by Zambian staff also contributed to cost-savings: notably, annual program cost per hospital was lower than SPINZ due to the exclusion of US-based staff salaries and travel expenses during the preparation period. Preparation costs were also notably lower, as it was assumed that after the first year of the program, only facility-based refresher trainings would occur (rather than full-day trainings requiring external venue hire and accommodation). There may also be opportunities for further cost-saving if this program were to be taken to scale, as the majority of costs are supplies, but there is possibly for lower prices to be negotiated if bulk-buying for a national program. The future estimate also accounts for brand new stock every year, but it is possible that this would not be needed for things that can be reused (e.g., wall-mounted dispensers for alcohol hand rub).

There are several limitations that should be noted. First, SPINZ was a single site observational study rather than a randomized controlled trial. While the results were robust and significant, some experts may prefer to see the results of a randomized trial (and possibly in multiple settings) before concluding the intervention is generalizable to other low- and middle-income countries. However, randomization at the individual level is not feasible when a bundle of new interventions is being tested, so such a study would ideally be designed as a cluster randomized, controlled trial with NICUs serving as the clusters [23]. The cost of such a study would be substantial. Second, the timeframe of delivering the intervention was relatively short. That said, the intervention was provided for almost 12 continuous months, thus allowing for the effect of seasonal changes. Third, we did not conduct a prospective, micro-costing economic analysis, which is considered a more precise costing method for these types of studies, since the analysis was a secondary, follow-on endeavor. Fourth, our estimates of future costs are extrapolated from a single institution. Not all district hospitals have NICUs or special care nurseries with the same level of care available in the UTH NICU and other NICUs may have different neonatal mortality rates. Finally, the SPINZ study focused on neonates who had nosocomial infections (after at least 2 days in the NICU and only included those whose parents provided informed consent. While the majority of babies admitted to the NICU participated in the study, neonatal deaths in the first 48 hours were excluded from the analysis. Since these newborns might also benefit from the bundle of interventions, the effect might be even greater in terms of benefit and cost effectiveness.

## Conclusion

The multi-component bundle of IPC interventions used in the SPINZ trial was a highly cost-effective strategy for reducing nosocomial BSI and neonatal mortality. Alcohol-based hand rub is an important part of such interventions although behavior change communication is also a vital component. We recommend scale-up of the SPINZ IPC bundle where neonatal sepsis and other nosocomial infection rates are high, as well as future research to confirm the impact and cost effectiveness of IPC bundles in different contexts in LMICs, including at health care facilities at lower levels of the health system.

## Data Availability

Due to restrictions on sharing data externally imposed by the National Health Authority of Zambia, requests for a limited data set for the clinical data must be made to the Zambia study team and then a data sharing agreement can be initiated. Requests for the Excel-based cost data can be directed to the first author and will be provided on request.

## Acknowledgements

We would like to thank the SPINZ study team including James Mwansa, Chileshe Lukwesa-Musyani, Angela Lyondo, Moses Chilufya Malama, Russell Localio, Cassandra Pierre, Surabhi Bhatt, Gertrude Munanjala, Nellisiwe Chizuni, Aja Griffin, and Caitryn McCallum for their contributions.

## Supporting information

**S1 Table. Pre-intervention demographics and outcomes for SPINZ study by month.**

**S2 Table. Demographics and outcomes for SPINZ study by intervention month.**

## Notes

### Competing Interest Statement

The authors have declared no competing interest.

### Clinical Trial

NCT02386592

### Funding Statement

The SPINZ study was funded by the Thrasher Research Fund (grant #12036). The study was designed, analyzed and implemented by the authors. The funder had no role in data collection, analysis, or interpretation.

## References

[1] Hug L, Alexander M, You D, et al. National, regional, and global levels and trends in neonatal mortality between 1990 and 2017, with scenario-based projections to 2030: a systematic analysis. Lancet Glob Health 2019; 7: e710–e720.

[2] Bhutta ZA, Das JK, Bahl R, et al. Can available interventions end preventable deaths in mothers, newborn babies, and stillbirths, and at what cost? The Lancet 2014; 384: 347–370.

[3] Fleischmann-Struzek C, Goldfarb DM, Schlattmann P, et al. The global burden of paediatric and neonatal sepsis: a systematic review. Lancet Respir Med 2018; 6: 223–230.

[4] Reinhart K, Schachter RD. Recognizing sepsis as a global health priority — A WHO resolution. N Engl J Med 2017; 377: 414–417.

[5] Molloy EJ, Bearer CF. Paediatric and neonatal sepsis and inflammation. Pediatr Res 2022; 91: 267–269.

[6] Celik IH, Hanna M, Canpolat FE, et al. Diagnosis of neonatal sepsis: the past, present and future. Pediatr Res 2022; 91: 337–350.

[7] Newborn mortality. World Health Organization, https://www.who.int/news-room/fact-sheets/detail/newborn-mortality#:~:text=Overview,in%20child%20survival%20since%201990. (25 March 2024, accessed 1 June 2025).

[8] Doctor HV, Nkhana-Salimu S, Abdulsalam-Anibilowo M. Health facility delivery in sub-Saharan Africa: successes, challenges, and implications for the 2030 development agenda. BMC Public Health 2018; 18: 765.

[9] Moyer CA, Mustafa A. Drivers and deterrents of facility delivery in sub-Saharan Africa: a systematic review. Reprod Health 2013; 10: 40.

[10] Zaidi AK, Huskins WC, Thaver D, et al. Hospital-acquired neonatal infections in developing countries. The Lancet 2005; 365: 1175–1188.

[11] Amare D, Mela M, Dessie G. Unfinished agenda of the neonates in developing countries: magnitude of neonatal sepsis: systematic review and meta-analysis. Heliyon 2019; 5: e02519.

[12] Kamanga A, Ngosa L, Aladesanmi O, et al. Reducing maternal and neonatal mortality through integrated and sustainability-focused programming in Zambia. PLOS Glob Public Health 2022; 2: e0001162.

[13] Zambia Demographic and Health Survey 2018. Lusaka, Zambia: Zambia Statistics Agency, Ministry of Health, https://dhsprogram.com/publications/publication-fr361-dhs-final-reports.cfm (2019, accessed 15 April 2023).

[14] Mwananyanda L, Pierre C, Mwansa J, et al. Preventing bloodstream infections and death in Zambian neonates: Impact of a low-cost infection control bundle. Clin Infect Dis 2019; 69: 1360–1367.

[15] Sabin LL, Knapp AB, MacLeod WB, et al. Costs and cost-effectiveness of training traditional birth attendants to reduce neonatal mortality in the Lufwanyama Neonatal Survival Study (LUNESP). PLoS ONE 2012; 7: e35560.

[16] Biemba G, et al. Newborn care in Zambia: A country case study of facility-based maternal and newborn care.

[17] Zulu J, Mwansa G, Changwe K. Forecasting inflation rate using the ARIMA model: Zambia’s perspective from 2023 to 2043. Int J Res Sci Innov 2025; XI: 698–713.

[18] Attema AE, Brouwer WBF, Claxton K. Discounting in economic evaluations. PharmacoEconomics 2018; 36: 745–758.

[19] Murray CJL. Quantifying the burden of disease: the technical basis for disability-adjusted life years. Bull World Health Organ 1994; 72: 429–445.

[20] Sachs J (ed). Macroeconomics and health: investing in health for economic development ; report of the Commission on Macroeconomics and Health. Geneva: World Health Organization, 2001.

[21] Lee Him R, Rehman S, Sihota D, et al. Prevention and treatment of neonatal infections in facility and community settings of low- and middle-income countries: A descriptive review. Neonatology 2024; 1–36.

[22] Fenny AP, Otieku E, Labi KA-K, et al. Cost-effectiveness analysis of alcohol handrub for the prevention of neonatal bloodstream infections: Evidence from HAI-Ghana study. PLOS ONE 2022; 17: e0264905.

[23] Him R, Sihota D, Harrison L, et al. Strategies to reduce antimicrobial resistance in newborns in low- and middle-income countries: a systematic review and meta-analysis. Lancet Glob Health.

